# Racial Disparities in Adverse Cardiovascular Events After Cardiac Electrophysiology Procedures: A Real-World, Multi-Institutional Retrospective Cohort Study

**DOI:** 10.1101/2025.02.17.25322440

**Authors:** Ugochukwu Ebubechukwu, Chinenye Okafor, Anderson Anuforo, Onyinye Ugoala, Hassan Tariq, Sana Dang, Azka Naeem, Joseph E. Marine, Festus Ibe, Kamala P. Tamirisa

## Abstract

**Background:** Electrophysiology (EP) procedures have revolutionized the management of arrhythmias, improving survival and overall outcomes. However, limited data exists evaluating the long-term outcomes of different EP procedures in underrepresented racial and ethnic groups.

**Methods:** We utilized data from the TriNetX US collaborative network and included adult participants who underwent EP procedures between 2013-2023 to create two cohorts: non-Hispanic Black and non-Hispanic White participants. Propensity score matching (PSM) was performed using predefined sociodemographic characteristics, medical comorbidities and medications used to ensure balance between groups. Primary outcomes were all-cause mortality at 30-days and 1-year post-EP procedures. Secondary outcomes included new-onset acute myocardial infarction (AMI), ischemic stroke, 3-point major adverse cardiovascular events (MACE), cardiac arrest, ventricular fibrillation (V-fib), ventricular tachycardia (VT), heart failure (HF), and procedure complications including pneumothorax and cardiac tamponade during follow-up.

**Results:** After PSM, we had improved balance between cohorts with 69,620 matched participants: 50.0% non-Hispanic Black participants with a mean age of 60.7 years; 43.6% females. Compared to non-Hispanic White participants, non-Hispanic Black participants had similar risk of all-cause mortality at 30-days (HR 0.94 95% CI; 0.84,1.06), and 1-year (HR 1.02 95% CI; 0.96,1.08) post-EP procedure after PSM. Moreover, non-Hispanic Black participants had significantly higher risk of developing AMI (HR 1.25 95% CI; 1.10,1.41), 3-point MACE (HR 1.15 95% CI; 1.04,1.27), VT (HR 1.29 95% CI; 1.18,1.42), V-fib (HR 1.28 95% CI; 1.09,1.51) and HF (HR 1.33 95% CI; 1.24,1.42) at 30 days, and these findings remained consistently significant at 1-year follow-up after PSM. No significant differences were noted in all other studied outcomes.

**Conclusion:** Non-Hispanic Black patients who underwent any EP procedure had a similar risk of 30-day, and 1-year mortality when compared to non-Hispanic White patients. However, non-Hispanic Black participants had higher rates of adverse cardiovascular events in the short and long-term than non-Hispanic White participants post-EP procedure.

**Clinical Perspective:** *What is New?:*

- This multi-institutional study addresses the question: Are the odds of developing an adverse cardiovascular outcome following an EP procedure the same for a non-Hispanic Black patient when compared to a non-Hispanic White patient?
- Non-Hispanic Black patients who underwent any EP procedure had a similar risk of 30-day, and 1-year mortality when compared to non-Hispanic White patients. However, non-Hispanic Black patients showed a statistically significant higher prevalence of adverse cardiovascular outcomes such as 3-point major adverse cardiovascular events (MACE), acute myocardial infarction, ventricular tachycardia, ventricular fibrillation, and heart failure.

*What are the clinical implications?:*

- This study addresses the racial differences in short– and long-term adverse cardiovascular events following an EP procedure. Better understanding of factors (such as socioeconomic status, health literacy, environmental circumstances etc.) that could potentially limit access to healthcare access among underrepresented racial and ethnic groups (UREG) will better inform policymakers to implement strategies to mitigate these differences.

## Introduction

Cardiovascular diseases (CVD) continue to be the leading cause of death in the United States.^1^ Among the various manifestations, cardiac arrhythmias constitute a significant proportion. Specialized electrophysiology (EP) interventions including catheter ablation, pacemaker implantation, and implantable cardioverter-defibrillator (ICD) placement have revolutionized the management of arrhythmias and contributed to improved survival outcomes.^2^ Though the past decades have witnessed a decline in cardiovascular mortality rates, these benefits have not been equitably distributed across various racial and ethnic groups.^3^

For example, Black patients hospitalized with heart failure who are eligible for ICD placement have significantly lower odds of receiving an ICD than White patients.^4^ Additionally, Black patients are less likely to undergo catheter ablation for ventricular arrhythmias and have higher risk-adjusted 1-year mortality rates after hospital discharge compared to White counterparts.^5^ Disparities in outcomes have been documented in patients undergoing cardiac resynchronization therapy, with Black and Hispanic patients experiencing higher odds of left ventricular lead implant failure.^6^ Similarly, racial gap has also been observed in the utilization of left atrial appendage occlusion (LAAO).^7^

Despite these concerning trends, limited research exists addressing racial disparities in post-EP procedures. Most studies focus on the immediate post-recovery period, leaving a gap in understanding the long-term outcomes in underrepresented racial and ethnic groups (UREG). To address these gaps, this study aims to examine the differences in all-cause mortality at 30-days, and 1-year following cardiac EP procedures among non-Hispanic Black participants versus non-Hispanic White participants. We also aim to evaluate adverse cardiovascular events among non-Hispanic Black participants when compared with non-Hispanic White participants at 30-days, and 1-year post-EP procedures. We hypothesize that non-Hispanic Black participants will experience worse outcomes, including higher rates of all-cause mortality and adverse cardiovascular events at all defined timepoints post-EP procedures when compared to non-Hispanic White participants.

## Methods

### Data source and collection

We performed a retrospective cohort study utilizing data from the TriNetX global health research within the US Collaborative Network. TriNetX is a global federated health research network providing access to de-identified data from electronic medical records (diagnoses, procedures, medications, laboratory values, genomic information) of more than 117 million patients across large healthcare organizations (HCOs) in the United States.^8^ This network included 67 HCO(s). The HCOs contributing de-identified electronic medical records (EMR) data include academic university hospitals, community hospitals, and specialty physician services providing data from inpatient and outpatient encounters. TriNetX complies with the Health Insurance Portability and Accountability Act (HIPAA), the US federal law, which protects the privacy and security of healthcare data. All data (either patient-level in a dataset or aggregate) exhibited on the TriNetX Platform only contains de-identified data per the de-identification standard defined in Section §164.514(a) of the HIPAA Privacy Rule.^8^ TriNetX federated network does not share patient-identifiable information for research studies and is waived from Western Institutional Review Board (IRB).^8^

### Study Population

Our study included adults (age **≥** 18 years old) participants who underwent any EP procedure between January 1, 2013, and December 31, 2023. Included participants were grouped into two cohorts based on race/ethnicity: Cohort A included non-Hispanic Black or African American participants (HL7V3.0:Race:2054-5 and Not Hispanic or Latino, HL7V3.0:Ethnicity:2186-5), and Cohort B included non-Hispanic White participants (White, HL7V3.0:Race:2106-3 and Not Hispanic or Latino, HL7V3.0:Ethnicity:2186-5). Patients were excluded, if (1) they were < 18 years old; (2) they were ≥ 90 years old; (3) patients that were neither non-Hispanic Black nor non-Hispanic White race. The included EP procedures comprised intracardiac catheter ablation for atrial fibrillation, supraventricular tachycardia, and ventricular tachycardia, along with intracardiac 3-D mapping, pacemaker implantation, pacemaker pulse generator replacement, pacemaker removal, cardiac resynchronization therapy, repositioning of previously implanted pacemaker, and implantable cardioverter-defibrillator implantation as defined by the International Classification of Diseases, Tenth Revision, Clinical Modification (ICD-10-CM) and Current Procedural Terminology (CPT) coding (**Table S1**). Sociodemographic information included race, biological sex, and age. We also collected information on co-existent medical conditions including essential hypertension, diabetes mellitus, obesity, chronic kidney disease, thyroid gland diseases, and obstructive sleep apnea (OSA). Additionally, we recorded medications used during the study period such as, diuretics, beta-blockers, calcium channel blockers, angiotensin-converting enzyme inhibitors (ACE-i), aldosterone receptor blockers (ARBs), statins, antiarrhythmics, antiplatelets, and anticoagulants.

The index event, defined as undergoing an EP procedure, marked as the point at which each patient entered the analysis. The time window was defined as the duration during which outcomes were analyzed. This analysis included outcomes that occurred within a time window starting on the day as the first EP procedure and extending to 30 days and one year post-procedure.

### Primary and Secondary outcomes

All outcome measures were defined using ICD-10-CM codes. Primary outcomes were all-cause mortality within 30-days and 1-year following any of the predefined EP procedures. Secondary outcome measures included new-onset acute myocardial infarction (AMI, ICD-10-CM:I21), ischemic stroke (ICD-10-CM:I63), and 3-point major adverse cardiovascular events (MACE), defined as nonfatal stroke, nonfatal myocardial infarction, and cardiovascular death, (ICD-10-CM:I21 and I63), cardiac arrest (ICD-10-CM:I46 or I97.12 or I97.71), ventricular fibrillation (V-fib, ICD-10-CM:I49.0), ventricular tachycardia (VT, ICD-10-CM:I47.2), heart failure (HF, ICD-10-CM:I50), and complications including pneumothorax (ICD-10-CM:J93), and cardiac tamponade (ICD-10-CM:I31.4) within 30 days and 1-year post EP procedure.

### Statistical analysis

All analyses for this study were conducted using the TriNetX platform. TriNetX has developed a data analysis platform that users can access directly on their website. We performed descriptive statistics on sociodemographic and clinical characteristics of participants, using counts and percentages for categorical variables and mean and standard deviation (SD) for continuous variables. Unadjusted logistic regression was used to determine factors associated with predefined outcomes between the two cohorts. The initial dataset included 35,100 non-Hispanic Black participants and 252,207 non-Hispanic White participants. See **Figure 1**.

**Figure 1:**
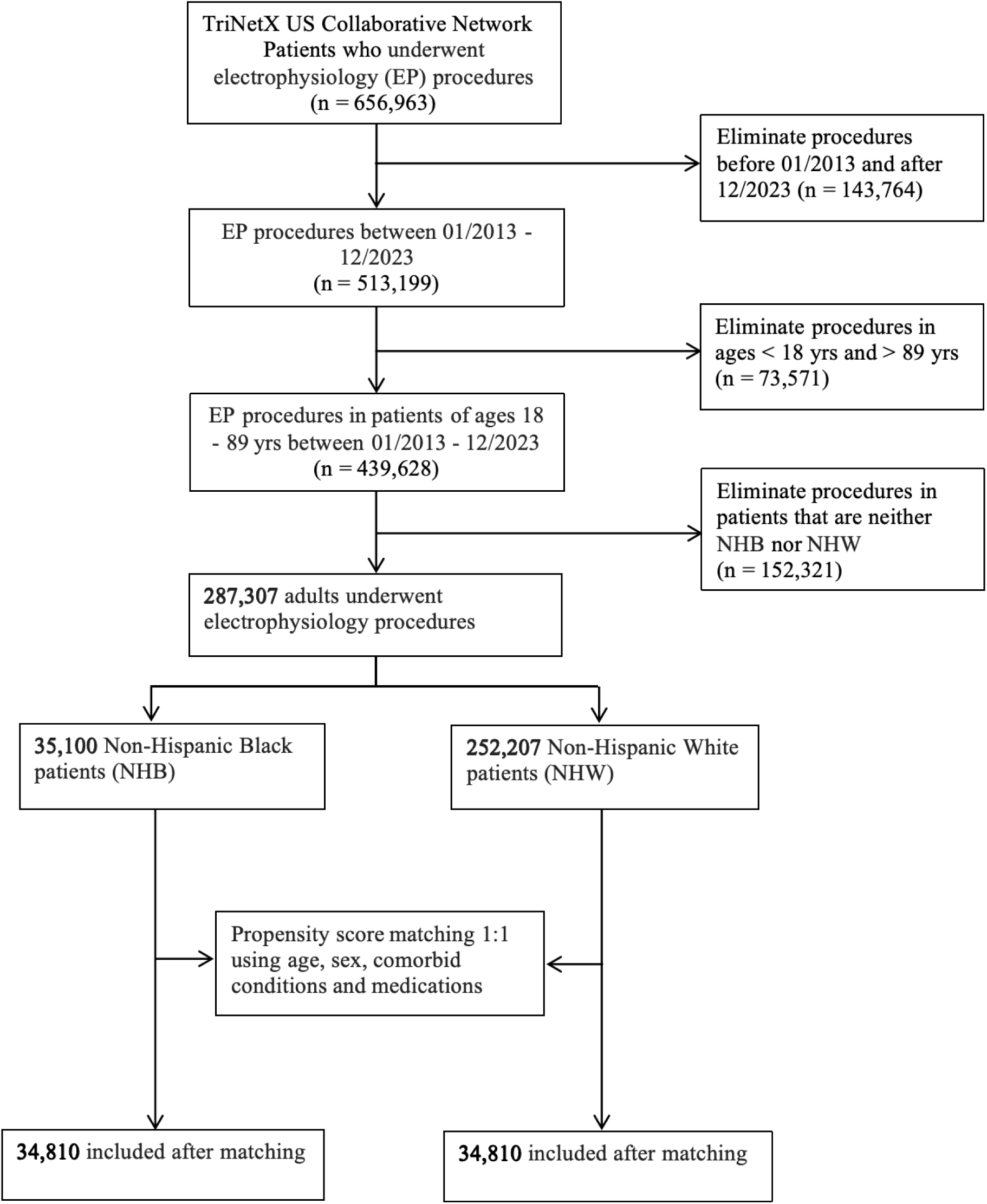
Study Flow Chart.

We subsequently conducted propensity score matching (PSM) between the two cohorts with a 1:1 greedy (nearest neighbor) matching structure to reduce confounding. PSM accounted for sociodemographic data (age, sex), co-existing conditions (hypertension, diabetes mellitus, obesity, chronic kidney disease, thyroid gland diseases, obstructive sleep apnea), and medications (diuretics, beta-blockers, calcium channel blockers, ACE-i/ARBs, statins, antiarrhythmics, antiplatelets and anticoagulants). PSM using 1:1 greedy (nearest neighbor) match on TriNetX has been utilized in prior research studies.^9–11^

After PSM, the balance of all included variables improved, as assessed using their standardized difference before and after matching (**Table 1**). Cox regression models on matched participants was used to calculate hazard ratios (HR) and 95% confidence intervals (CI) for all primary and secondary outcomes for non-Hispanic Black participants compared to non-Hispanic White participants. Kaplan-Meier curves were used to estimate the survival probability and log-rank test was used to determine significance in survival between the two cohorts. All analyses were conducted with significance set at p <0.05.

**Table 1.**
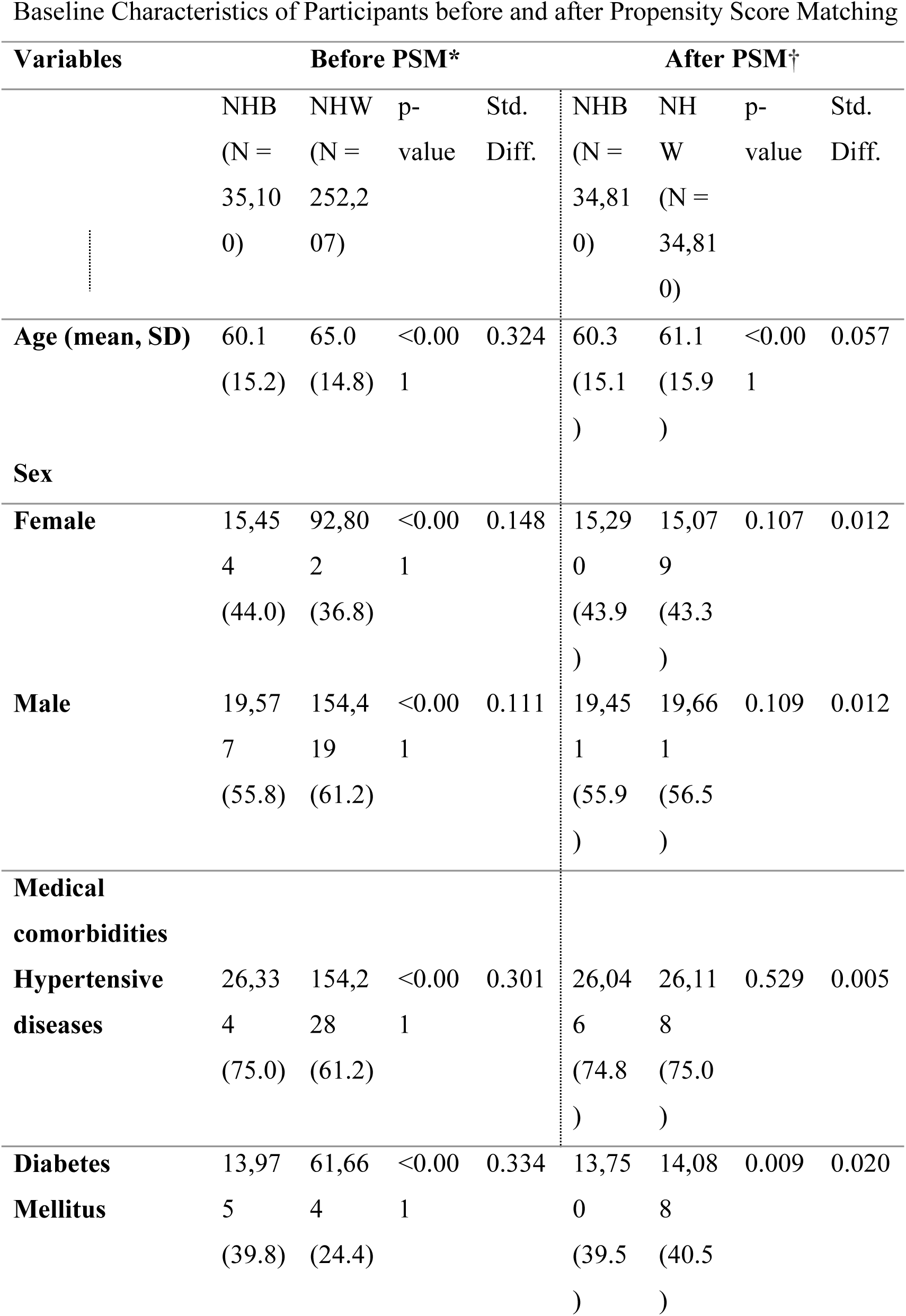
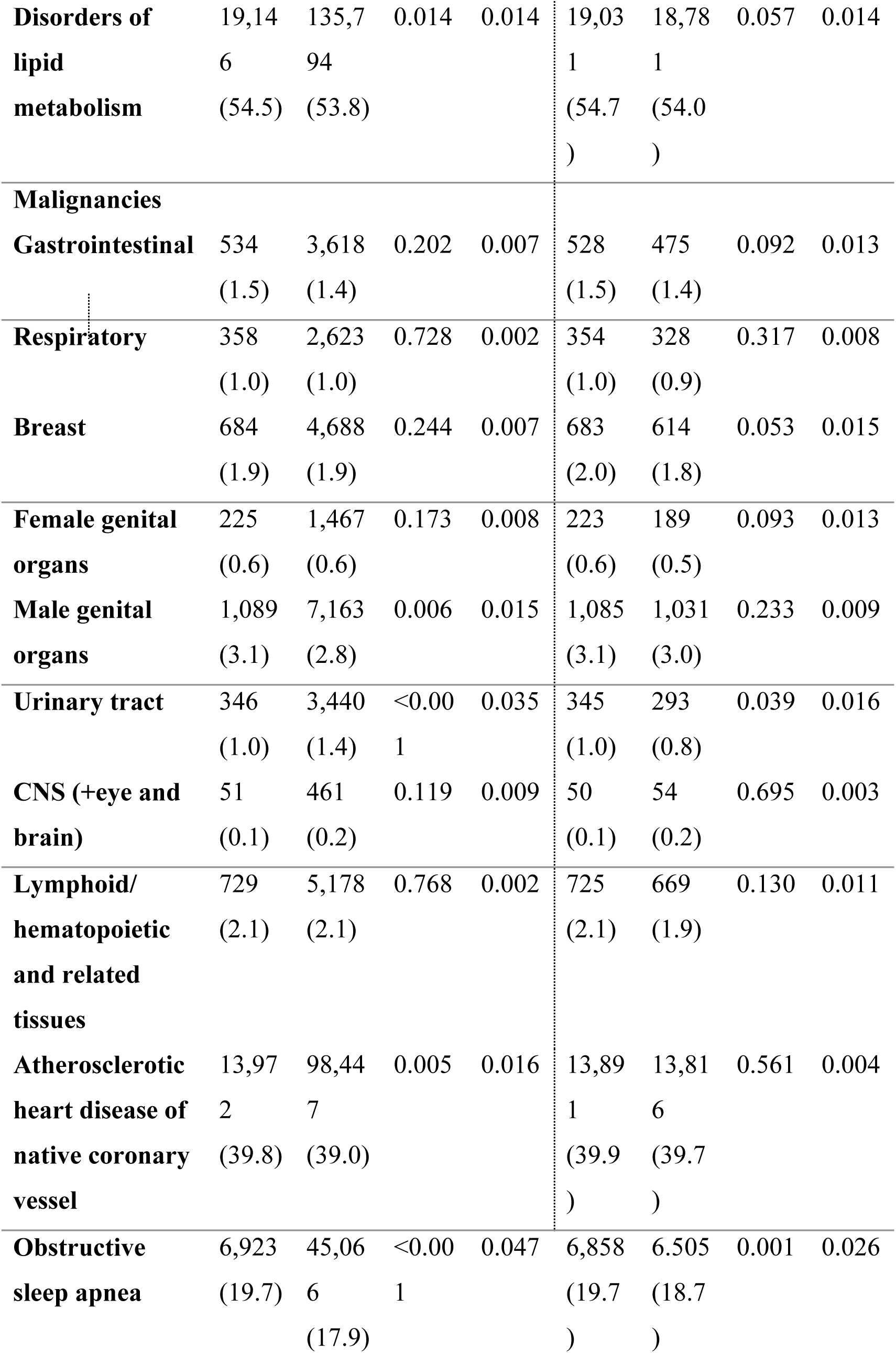
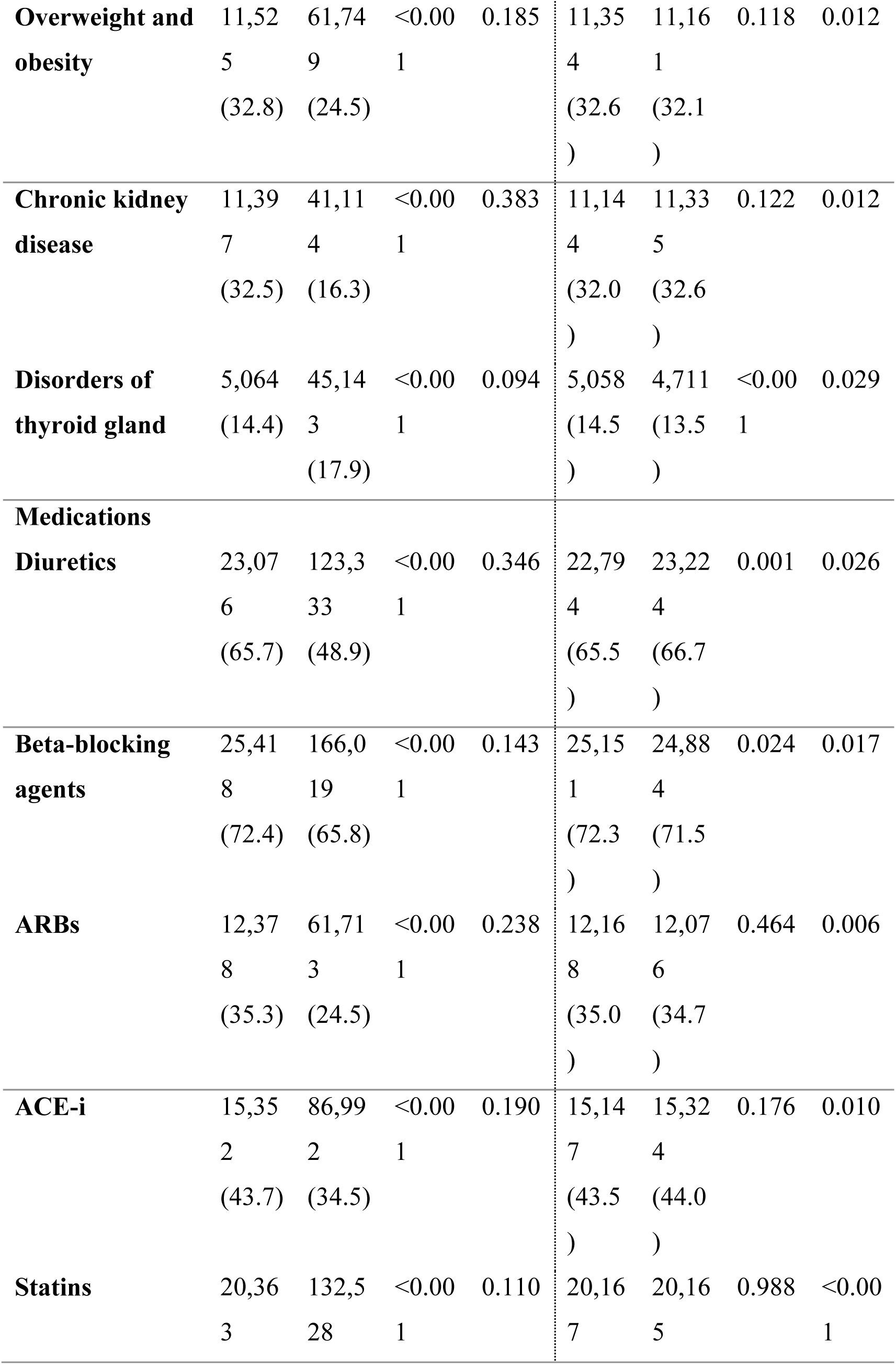
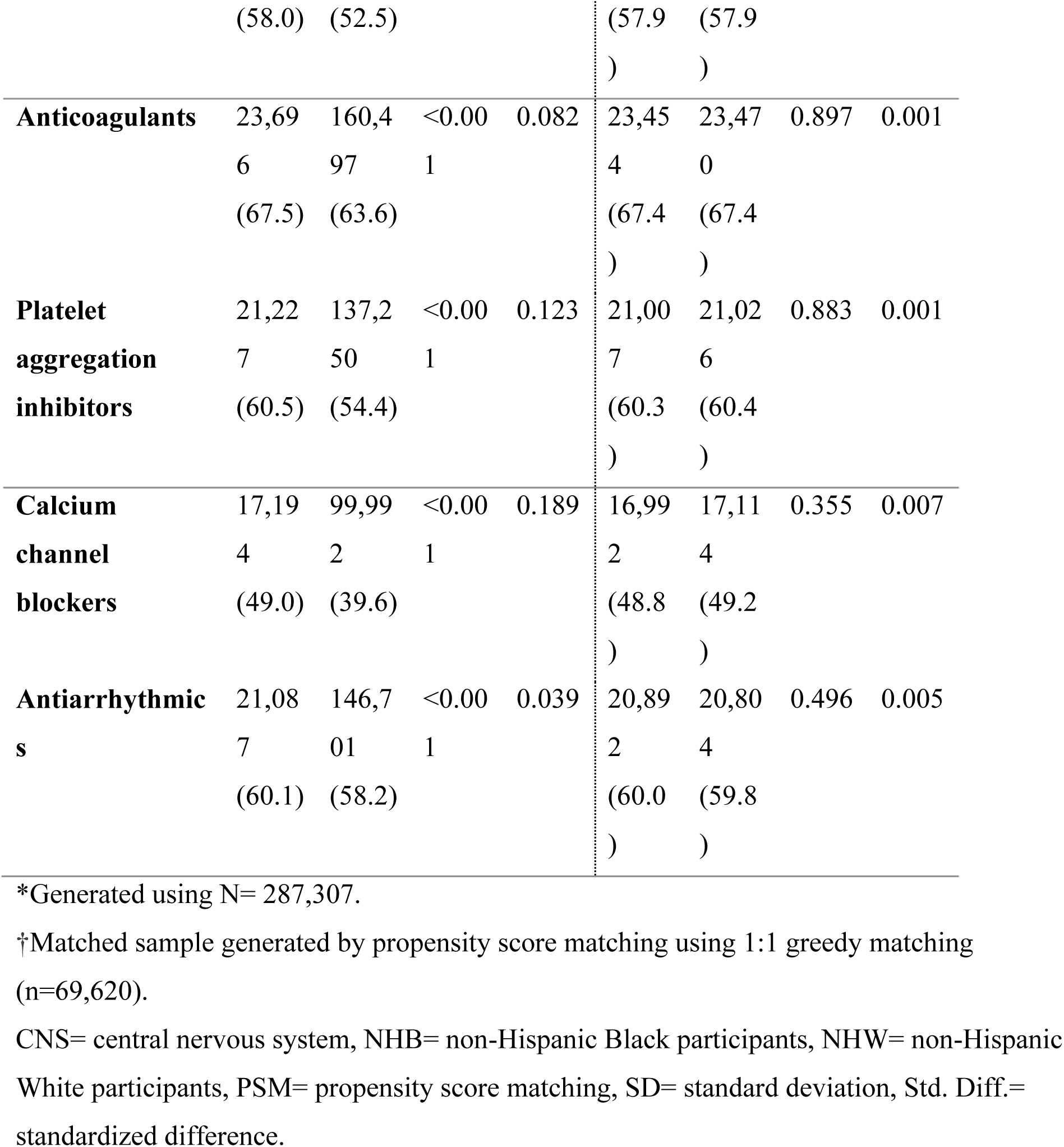
Baseline Characteristics of Participants before and after Propensity Score Matching.

This study was reported in compliance with the Strengthening the Reporting of Observational Studies in Epidemiology (STROBE) guidelines.^12^

## Results

### Baseline characteristics

Our study included 287,307 unmatched participants comprising 35,100 non-Hispanic Black participants and 252,207 non-Hispanic White participants. After PSM, we had 69,620 matched participants with 50.0% (34,810) being non-Hispanic Black participants, with a mean age of 60.7 years; 43.6% were females.

Before PSM, non-Hispanic Black participants were significantly younger (60.1 years vs 65.0 years, p<0.001), and had a higher proportion of females (44.0% vs 36.8%, p<0.001) compared to non-Hispanic White participants. They also had a significantly higher prevalence of hypertension, diabetes mellitus, lipid disorders, OSA, overweight/obesity, and chronic kidney disease compared to non-Hispanic White counterparts (**Table 1**). Additionally, a significantly higher proportion of non-Hispanic Black participants were on diuretics, beta blockers, ARBs, ACE-i, statins, anticoagulants, antiplatelets, calcium channel blockers, and antiarrhythmics (**Table 1**).

After PSM, the cohorts were well-balanced. However, non-Hispanic Black participants were significantly older (60.3 years vs 61.1 years, p<0.001), and a higher prevalence of OSA (19.7% vs 18.7%, p=0.001) and thyroid gland disease (14.5% vs 13.5%, p<0.001) compared to non-Hispanic White participants (**Table 1**).

### 30-day, and 1-year outcomes

The post-PSM analysis results for outcomes within 30-days, and 1-year post-EP procedures are presented in **Table 2**. The number of patients included and excluded in each cohort are presented in **Table 3**.

**Table 2.** Survival Analysis within 30-days, and 1-year post-EP among non-Hispanic Black versus non-Hispanic White participants.

| Survival Probability † |  |  |  |  |  |
| --- | --- | --- | --- | --- | --- |
|  | NHB (%) | NHW (%) | HR | 95% CI | p-value* |
| **30 Days Follow-up** |  |  |  |  |  |
| **All-cause mortality** | 98.42 | 98.32 | 0.94 | 0.84, 1.06 | 0.327 |
| **Acute MI** | 98.12 | 98.50 | 1.25 | 1.10, 1.41 | <0.001 |
| **Ischemic Stroke** | 99.40 | 99.51 | 1.21 | 0.98, 1.50 | 0.074 |
| **3-point MACE** | 96.75 | 97.18 | 1.15 | 1.04, 1.27 | 0.005 |
| **Cardiac Arrest** | 98.77 | 98.66 | 0.91 | 0.80, 1.05 | 0.197 |
| **Ventricular**<br>**Tachycardia** | 96.01 | 96.92 | 1.29 | 1.18, 1.42 | <0.001 |
| **Ventricular**<br>**Fibrillation** | 99.00 | 99.22 | 1.28 | 1.09, 1.51 | 0.003 |
| **Heart Failure** | 89.56 | 92.03 | 1.33 | 1.24, 1.42 | <0.001 |
| **Pneumothorax** | 99.32 | 99.25 | 0.91 | 0.76, 1.08 | 0.279 |
| **Cardiac Tamponade** | 99.72 | 99.70 | 0.94 | 0.71, 1.25 | 0.673 |
| **1- Year Follow-up** |  |  |  |  |  |
| **All-cause mortality** | 92.80 | 92.95 | 1.02 | 0.96, 1.08 | 0.534 |
| **Acute MI** | 95.10 | 96.37 | 1.35 | 1.24, 1.47 | <0.001 |
| **Ischemic Stroke** | 97.34 | 98.11 | 1.41 | 1.26, 1.57 | <0.001 |
| **3-point MACE** | 88.81 | 90.81 | 1.23 | 1.16, 1.30 | <0.001 |
| **Cardiac Arrest** | 97.28 | 97.82 | 1.22 | 1.11, 1.36 | <0.001 |
| **Ventricular**<br>**Tachycardia** | 90.07 | 92.87 | 1.41 | 1.33, 1.50 | <0.001 |
| **Ventricular**<br>**Fibrillation** | 97.71 | 98.47 | 1.49 | 1.33, 1.68 | <0.001 |
| **Heart Failure** | 82.26 | 86.11 | 1.31 | 1.24, 1.39 | <0.001 |
| **Pneumothorax** | 98.72 | 98.72 | 0.98 | 0.85, 1.12 | 0.746 |
| **Cardiac Tamponade** | 99.53 | 99.52 | 0.97 | 0.77, 1.21 | 0.760 |

†Matched sample generated by propensity score matching using 1:1 greedy matching (n=69,620). \*Log-rank p-value NHB= non-Hispanic Black participants, NHW= non-Hispanic White participants, HR= hazard ratio, CI= confidence interval, Acute MI= acute myocardial infarction, MACE= major adverse cardiovascular event.

**Table 3:** Number of Patients Included and Excluded for Each Outcome Per Cohort Group During The 30-Day, and 1-Year Follow Up Period.

| **Cohort** | **Patients in cohort** | **Patients with outcome** | **Patients excluded** |
| --- | --- | --- | --- |
| **All-cause mortality** |  |  |  |
| **NHB** | 34,803 | 533 | 34,270 |
| **NHW** | 34,866 | 568 | 34,298 |
| **Acute MI** |  |  |  |
| **NHB** | 28,555 | 525 | 28,030 |
| **NHW** | 29,110 | 431 | 28,679 |
| **Ischemic Stroke** |  |  |  |
| **NHB** | 31,407 | 183 | 31,224 |
| **NHW** | 32,402 | 156 | 32,246 |
| **3-point MACE** |  |  |  |
| **NHB** | 26,206 | 829 | 25,377 |
| **NHW** | 27,335 | 755 | 26,580 |
| **Cardiac Arrest** |  |  |  |
| **NHB** | 32,924 | 398 | 32,526 |
| **NHW** | 32,976 | 436 | 32,540 |
| **Ventricular Tachycardia** |  |  |  |
| **NHB** | 27,164 | 1,065 | 26,099 |
| **NHW** | 28,783 | 876 | 27,907 |
| **Ventricular fibrillation** |  |  |  |
| **NHB** | 33,330 | 330 | 33,000 |
| **NHW** | 33,588 | 260 | 33,328 |
| **Heart Failure** |  |  |  |
| **NHB** | 15,190 | 1,562 | 13,628 |
| **NHW** | 18,877 | 1,484 | 17,393 |
| **Pneumothorax** |  |  |  |
| **NHB** | 34,429 | 230 | 34,199 |
| **NHW** | 34,279 | 253 | 34,026 |
| **Cardiac Tamponade** |  |  |  |
| **NHB** | 34,793 | 95 | 34,698 |
| **NHW** | 34,775 | 101 | 34,674 |

(1 Year)
| **Cohort** | **Patients in cohort** | **Patients with outcome** | **Patients excluded** |
| --- | --- | --- | --- |
| **All-cause mortality** |  |  |  |
| **NHB** | 34,655 | 2252 | 32,403 |
| **NHW** | 34,695 | 2,241 | 32,454 |
| **Acute MI** |  |  |  |
| **NHB** | 28,427 | 1,255 | 27,172 |
| **NHW** | 28,907 | 963 | 27,944 |
| **Ischemic Stroke** |  |  |  |
| **NHB** | 31,263 | 731 | 30,532 |
| **NHW** | 32,244 | 544 | 31,700 |
| **3-point MACE** |  |  |  |
| **NHB** | 26,085 | 2,629 | 23,393 |
| **NHW** | 27,106 | 2,277 | 24,829 |
| **Cardiac Arrest** |  |  |  |
| **NHB** | 32,779 | 821 | 31,958 |
| **NHW** | 32,790 | 675 | 32,117 |
| **Ventricular Tachycardia** |  |  |  |
| **NHB** | 27,044 | 2,431 | 24,613 |
| **NHW** | 28,561 | 1,866 | 26,695 |
| **Ventricular fibrillation** |  |  |  |
| **NHB** | 33,181 | 694 | 32,487 |
| **NHW** | 33,441 | 474 | 32,967 |
| **Heart Failure** |  |  |  |
| **NHB** | 15,092 | 2,466 | 12,626 |
| **NHW** | 18,557 | 2,407 | 16,150 |
| **Pneumothorax** |  |  |  |
| **NHB** | 34,278 | 404 | 33,874 |
| **NHW** | 34,149 | 414 | 33,735 |
| **Cardiac Tamponade** |  |  |  |
| **NHB** | 34,644 | 152 | 34,492 |
| **NHW** | 34,614 | 158 | 34,456 |

NHB= non-Hispanic Black participants, NHW= non-Hispanic White participants, Acute MI= acute myocardial infarction, MACE= major adverse cardiovascular event.

#### All-Cause Mortality

Non-Hispanic Black participants had similar risk of 30-day (HR 0.94 95% CI; 0.84,1.06), and 1-year (HR 1.02 95% CI; 0.96,1.08) all-cause mortality following EP procedures when compared with non-Hispanic White participants.

#### Adverse Cardiovascular Events at 30 Days

The risks were significantly higher for all events except ischemic stroke, cardiac arrest, pneumothorax, and cardiac tamponade among non-Hispanic Black participants compared to non-Hispanic White participants. Specifically, non-Hispanic Black participants were significantly more likely to develop: AMI (HR 1.25 95% CI; 1.10,1.41), 3-point MACE (HR 1.15 95% CI; 1.04,1.27), VT (HR 1.29 95% CI; 1.18,1.42), V-fib (HR 1.28 95% CI; 1.09,1.51), and HF (HR 1.33 95% CI; 1.24,1.42) within 30-days after EP procedures compared with their non-Hispanic White counterparts after adjusting for age, biological sex, medical comorbidities and medications.

#### Adverse Cardiovascular Events at 1-Year

Non-Hispanic Black participants had significantly higher risks for all events except pneumothorax and cardiac tamponade, which remained similar between both cohorts. Non-Hispanic Black participants were significantly more likely to have an AMI (HR 1.35 95% CI; 1.24,1.47), ischemic stroke (HR 1.41 95% CI; 1.26,1.57), 3-point MACE (HR 1.23 95% CI; 1.16,1.30), cardiac arrest (HR 1.22 95% CI; 1.11,1.36), VT (HR 1.41 95% CI; 1.33,1.50), V-fib (HR 1.49 95% CI; 1.33,1.68), and HF (HR 1.31 95% CI; 1.24,1.39) compared with non-Hispanic White participants after adjusting for age, biological sex, medical comorbidities and medications.

#### Survival Probability at 30-days, and 1-year

Non-Hispanic Black participants had a similar survival probability for all-cause mortality at 30-days (98.42% vs 98.32%, p=0.33) and 1-year (92.80% vs 92.95%, p=0.53) following EP procedures compared to non-Hispanic White participants **(Table 2).**

At 30 days post-EP procedure, survival rates were similar, except for HF, where non-Hispanic Black participants had lower survival rates (89.56% vs 92.03%, log-rank *P* < 0.001).

By 1-year follow-up, the survival rates in non-Hispanic Black participants had slightly declined for AMI (95.10% vs 96.37%, log-rank *P* < 0.001), ischemic stroke (97.34% vs 98.11%, log-rank *P* < 0.001), 3-point MACE (88.81% vs 90.81%, log-rank *P* < 0.001), VT (90.07% vs 92.87%, log-rank *P* < 0.001), and V-Fib (97.71% vs 98.47%, log-rank *P* < 0.001). Similar to the 30-day follow-up, survival rates for non-Hispanic Black participants showed a continuous downward trend (82.26% vs 86.11%, log-rank *P* < 0.001) compared to their non-Hispanic White counterparts. **(Table 2)**.

## Discussion

To the best of the authors’ knowledge, this study is the first to comprehensively analyze adverse cardiovascular events following cardiac EP procedures among non-Hispanic Black versus non-Hispanic White participants. Our findings reveal that non-Hispanic Black participants faced similar risk of all-cause mortality during follow-up at 30 days, and 1-year post-EP procedures. However, they exhibited an increased risk of AMI, 3-point MACE, V-fib and HF within 30 days, when compared to their non-Hispanic White counterparts. These disparities, observed in the immediate follow-up period, persisted during long-term follow-up at 1-year. The observed racial differences highlight the need for long-term follow-up post EP-procedures.

Our study uniquely examines the outcomes of multiple EP procedures across a diverse patient population, distinguishing it from prior studies that focused on isolated outcomes. For example, a prospective study by Kong et al., found that Black patients undergoing catheter ablation for ventricular tachycardia experienced higher rates of VT recurrence at 7 months, though these differences disappeared after multivariable adjustment.^13^ In contrast, our study demonstrated persistently higher risk of VT at 1-year among Non-Hispanic Black participants, even after adjusting for possible confounders. Another study reported a threefold higher likelihood of arrhythmic events in non-Hispanic Black patients undergoing EP-guided therapy^14^, consistent with our observation of elevated risk of VT and V-fib in this population within 30 days post-procedure. The exact reason for this outcome is unclear; however, several theories exist. One possibility is an increased susceptibility of non-Hispanic Black participants due to ethnic dissimilarity and variations in genetic makeup influencing EP responses. Additionally, disproportionate differences in structural remodeling, including a higher prevalence of left ventricular hypertrophy in Black patients, predisposing to potentially lethal arrhythmias, including proarrhythmic responses.^14^

Studies on ICD outcomes further highlight racial disparities. Akhabue et al found that Black and Hispanic patients experienced increased risk of death following ICD placement, particularly during hospitalization.^15^ However, Pokorney et al. showed that there was no significant interaction between race and mortality risk with ICD placement.^16^ Another study assessing outcomes in African Americans undergoing ICD implantation, showed an increased risk for all-cause death, especially during the first 2-years of follow-up^17^, aligning with our findings. Conversely, Judith et al., demonstrated equally reduced mortality in both African Americans and White patients receiving ICD therapy compared to placebo.^18^ Similarly, while cardiac resynchronization therapy was associated with reduced 24-month mortality across all racial groups in some studies, others reported higher cumulative probabilities of HF or death among Black patients after three years compared to White patients.

Racial and ethnic differences in cardiovascular care and outcomes have been well-documented across the U.S. healthcare system.^19^ The reasons for these differences in outcomes, remain incompletely understood and likely stem from a combination of patient, physician, access, and systemic factors. These differences cannot be fully explained by variations in disease processes alone, as our study demonstrates a persistent difference in outcomes between non-Hispanic Black and non-Hispanic White participants, even after accounting for sociodemographic characteristics, co-existing medical conditions, and medication use.

As the first study to comprehensively evaluate the post-EP procedure outcomes in UREG populations, our findings highlight the urgent need for further research to close the gaps in outcomes. Identifying differences in survival and adverse outcomes is crucial for understanding and addressing systemic inequities in healthcare access and delivery.^20^ While short-term outcomes, such as procedural success and immediate complications are essential data, future studies could focus on long-term effectiveness and impact of EP procedures, especially in specific patient populations. Chronic diseases including diabetes, hypertension, obesity, OSA and CKD that disproportionately affect UREG populations have cumulative effects on long-term procedural efficacy and outcomes. Similarly, structural factors such as socioeconomic status^21^, and quality of care^22^ have been shown to disproportionately affect UREG populations, may also play a vital role in overall gaps. A deeper understanding of potential confounding factors, disease trends, and longitudinal outcomes will better inform policymakers to develop targeted programs and interventions to mitigate these differences. **(Fig 2)**

**Figure 2.**
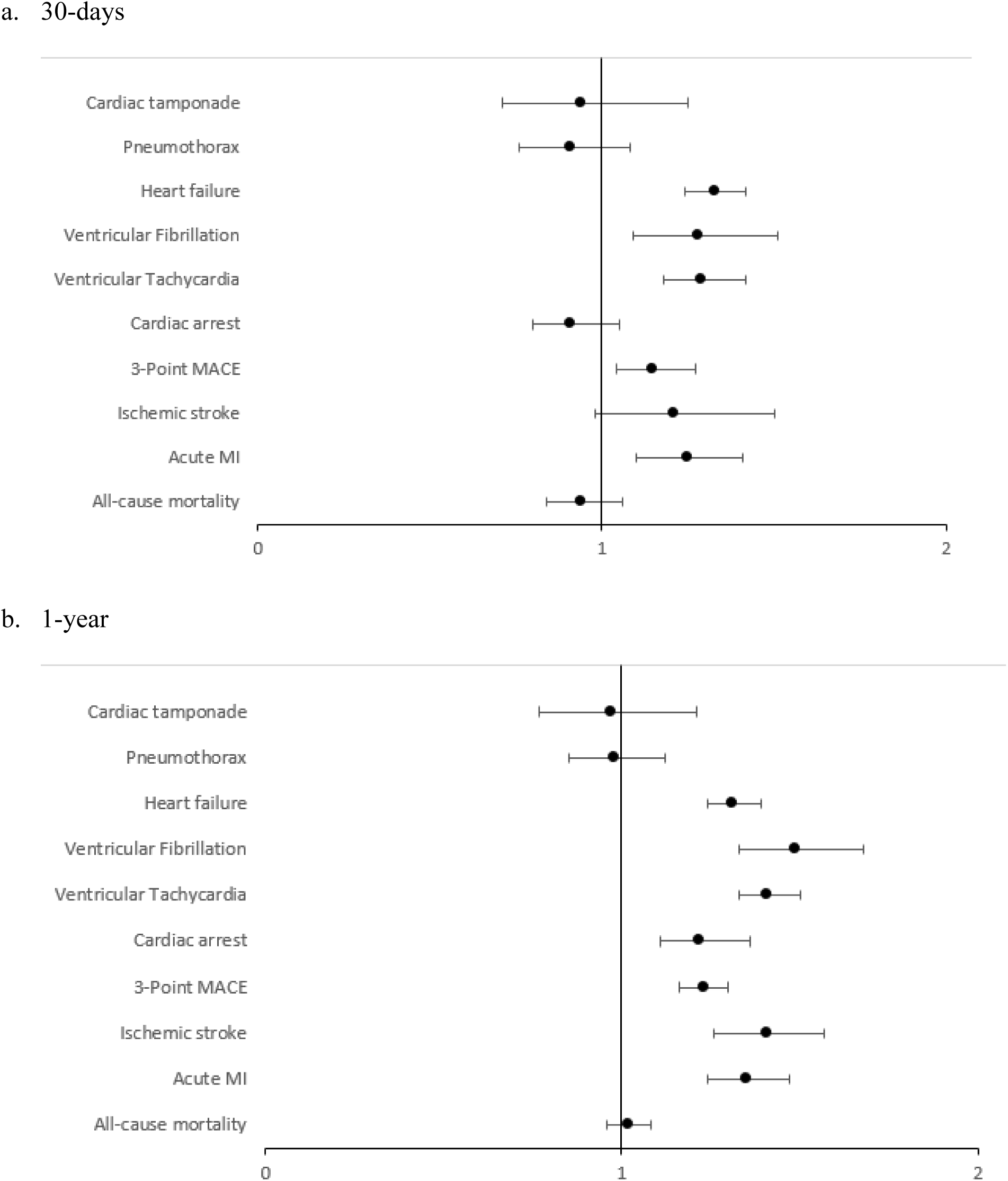
Forest plot for measured outcomes within 30-days, and 1-year post-EP among non-Hispanic Black versus non-Hispanic White participants.

**Figure 3.**
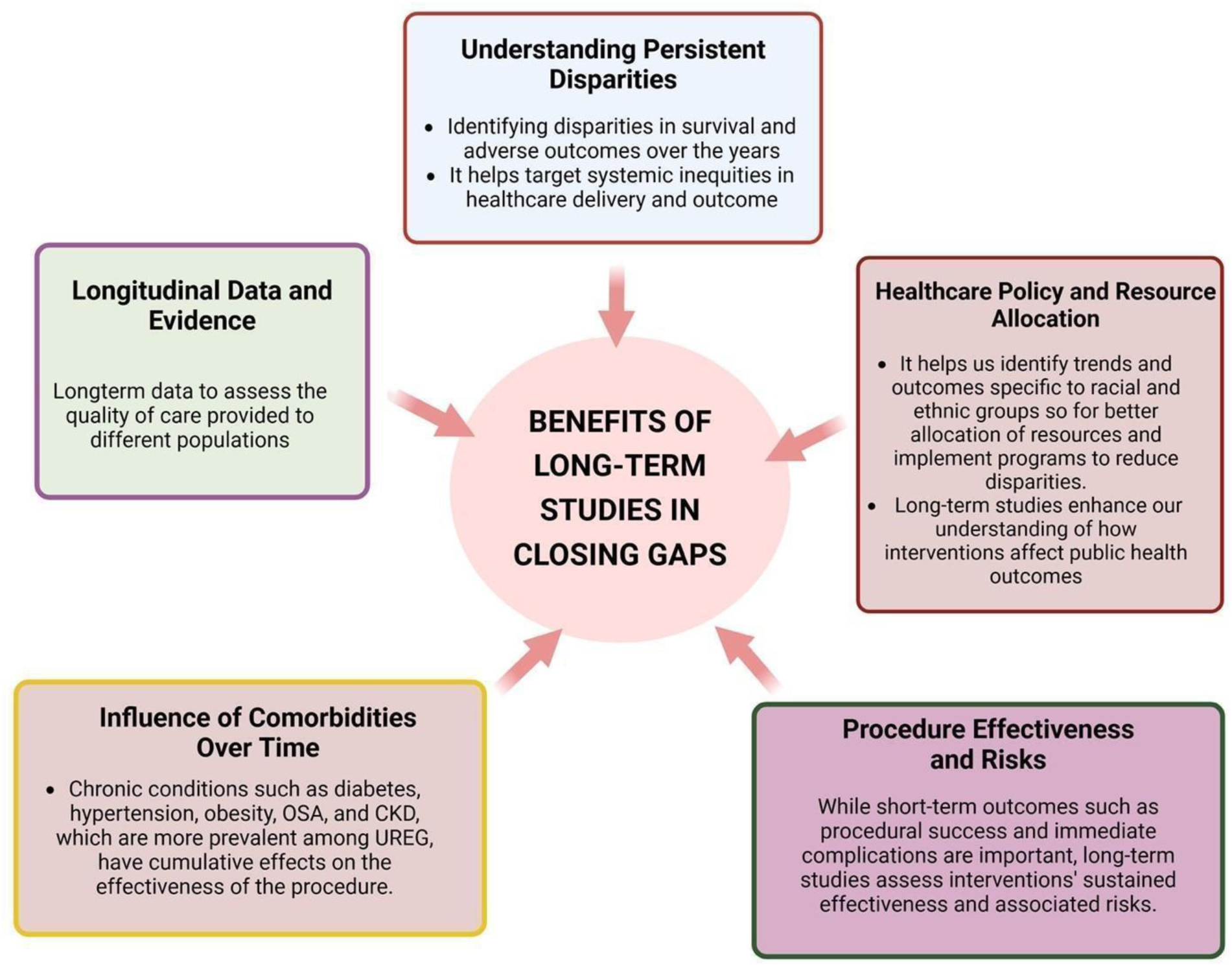
Benefits of long-term Studies in Closng Gaps.

## Conclusion

This retrospective cohort study provides a comprehensive analysis of short– and long-term outcomes following EP procedures, revealing that non-Hispanic Black participants face a higher risk of adverse cardiovascular events, including AMI, 3-point MACE, cardiac arrest, VT, V-fib and HF, at both 30-days and 1-year post procedure. Notably, these differences persisted even after adjusting for sociodemographic characteristics, medical comorbidities, and medications use, suggesting that factors beyond co-exist clinical conditions may contribute to these disparities.

## Limitations

Firstly, despite the utilization of a real-world database and detailed propensity score matching, this study faced certain limitations associated with data derived from EMR. These limitations included variability in provider data entry, challenges related to coding and diagnosis accuracy, and limited data on socioeconomic factors. Furthermore, as the data were sourced exclusively from HCOs within TriNetX network, outcomes occurring outside this database may not have been captured although this study ensured a comprehensive representation of the studied patient population by including the data from 67 large HCOs.

Although efforts were made to include all the commonly performed EP procedures, some, such as left atrial appendage occlusion procedures, were excluded due to their overlap with structural heart interventions. Given that our study focused on composite EP procedures, outcomes were not stratified by arrhythmia type and/or specific procedures. Additionally, while patients in each cohort were extensively matched, residual confounding factors related to medical history, patient sociodemographic characteristics, and/or procedure timing may have contributed to the observed differences in outcomes.

## Data source

The data supporting the findings of this study are available from the TriNetX Analytics Network. https://trinetx.com

## Nonstandard Abbreviations and Acronyms

CVD: Cardiovascular diseases
EP: Electrophysiology
HCO: Healthcare organization
NHB: Non-Hispanic Black
NHW: Non-Hispanic White
PSM: Propensity score matching
UREG: Underrepresented Racial and Ethnic Groups

---

**Table S1.**
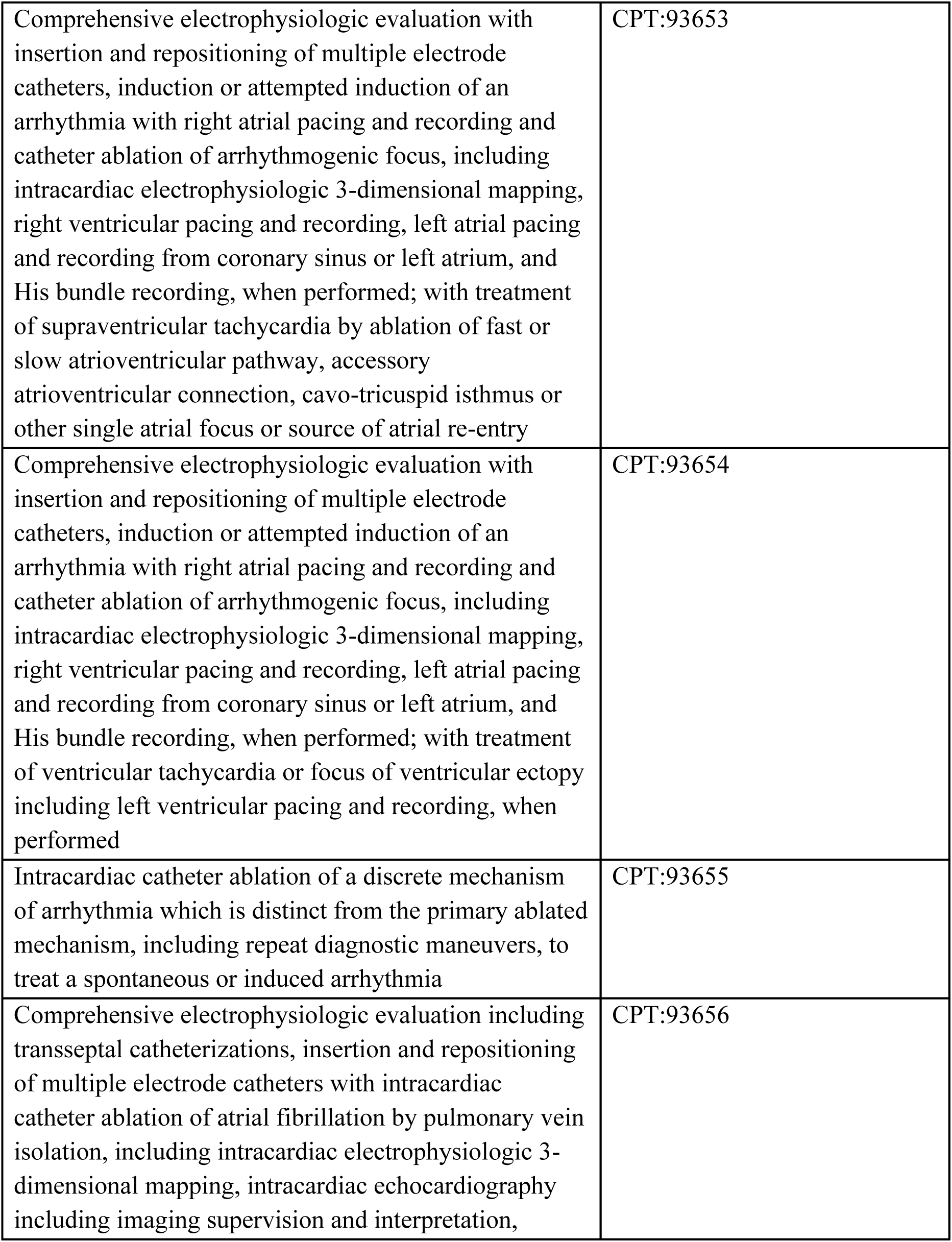
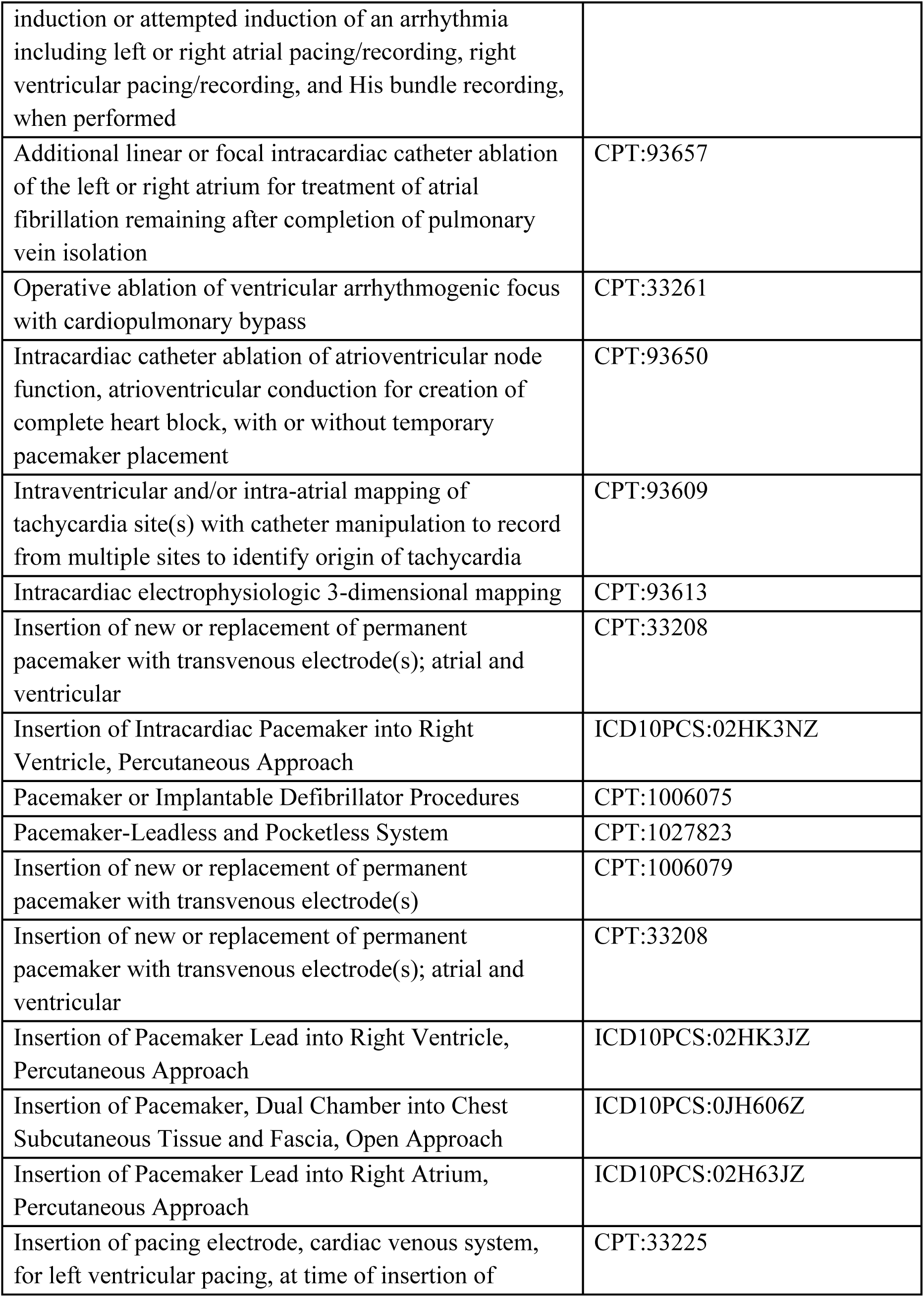
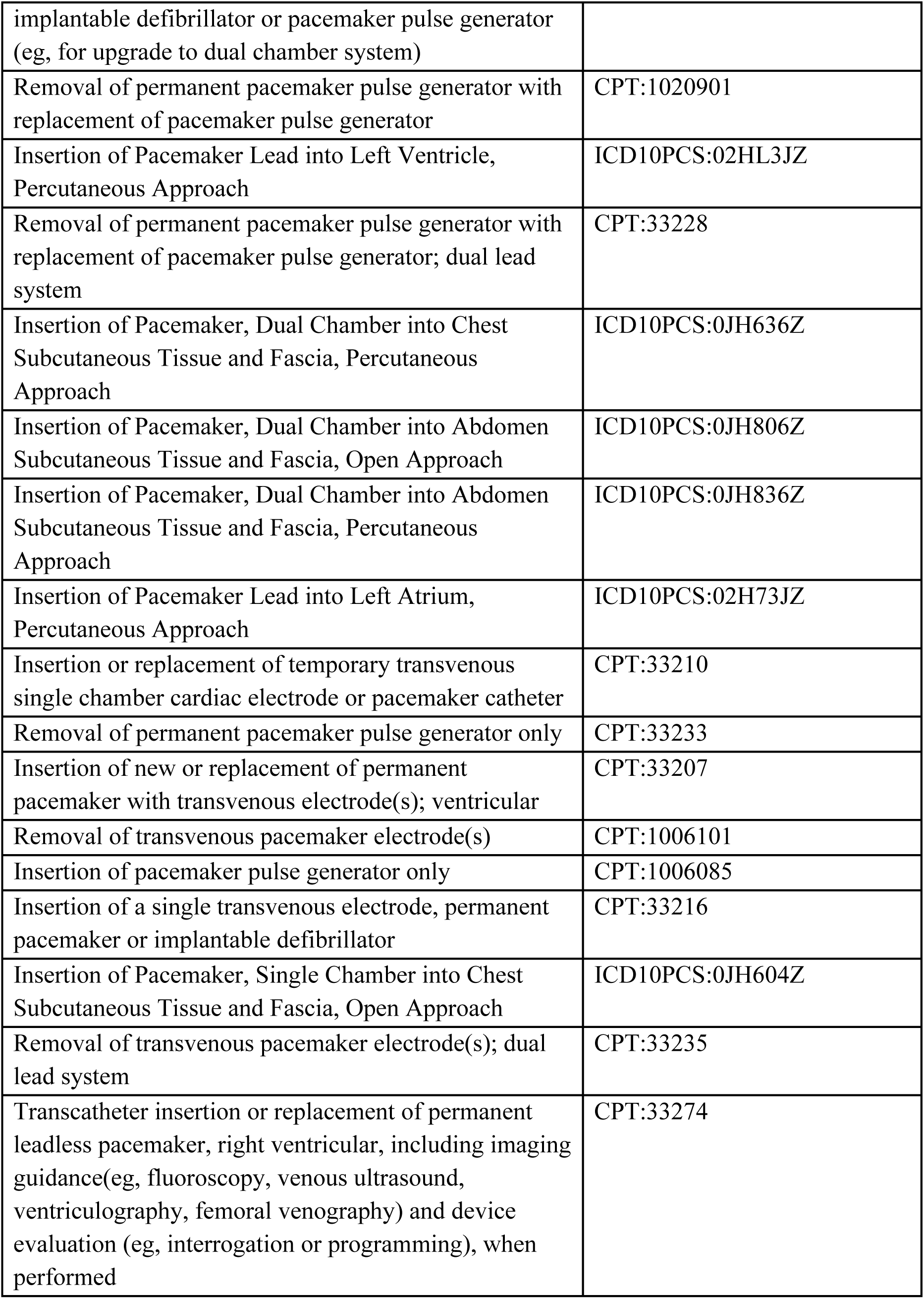
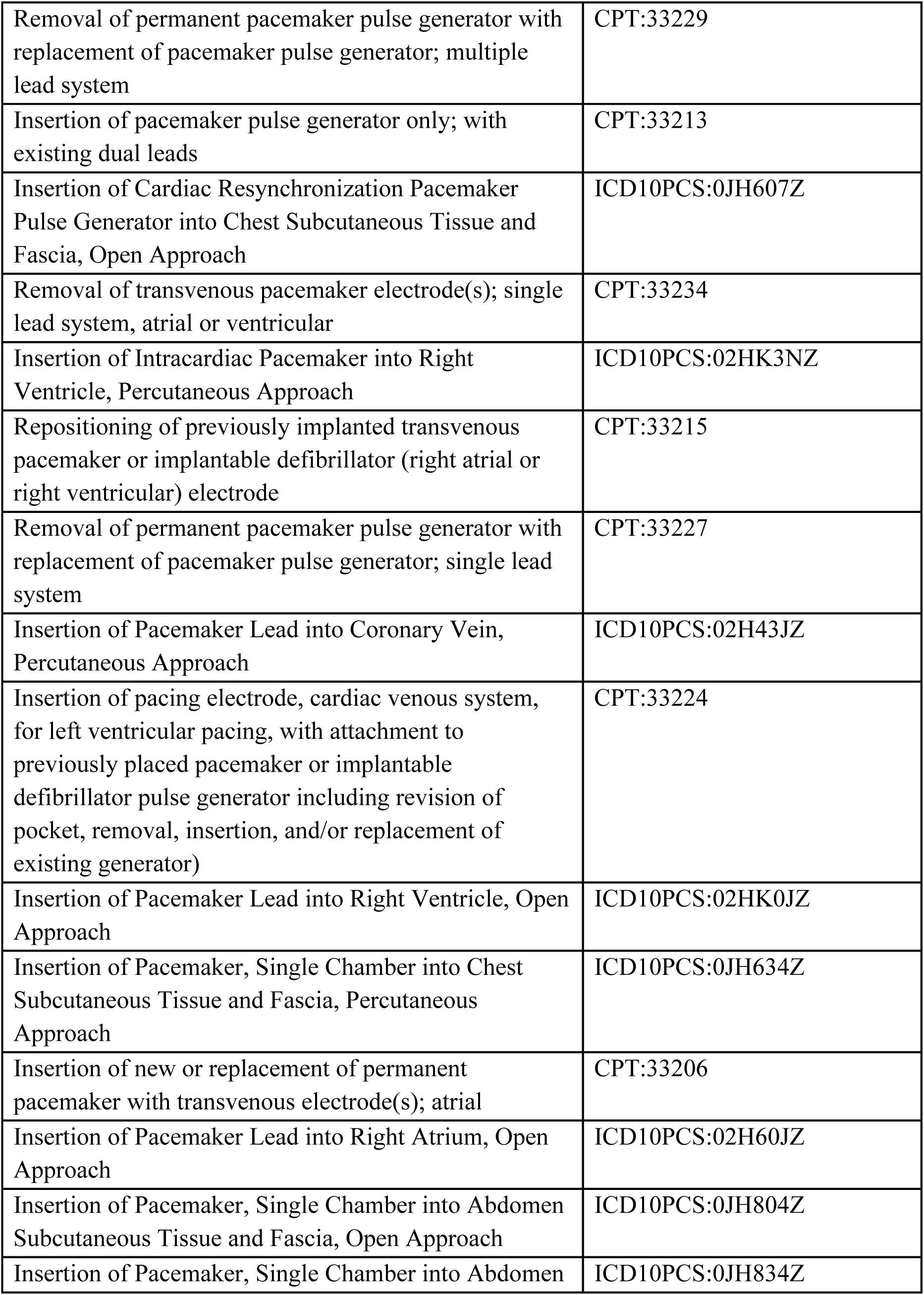
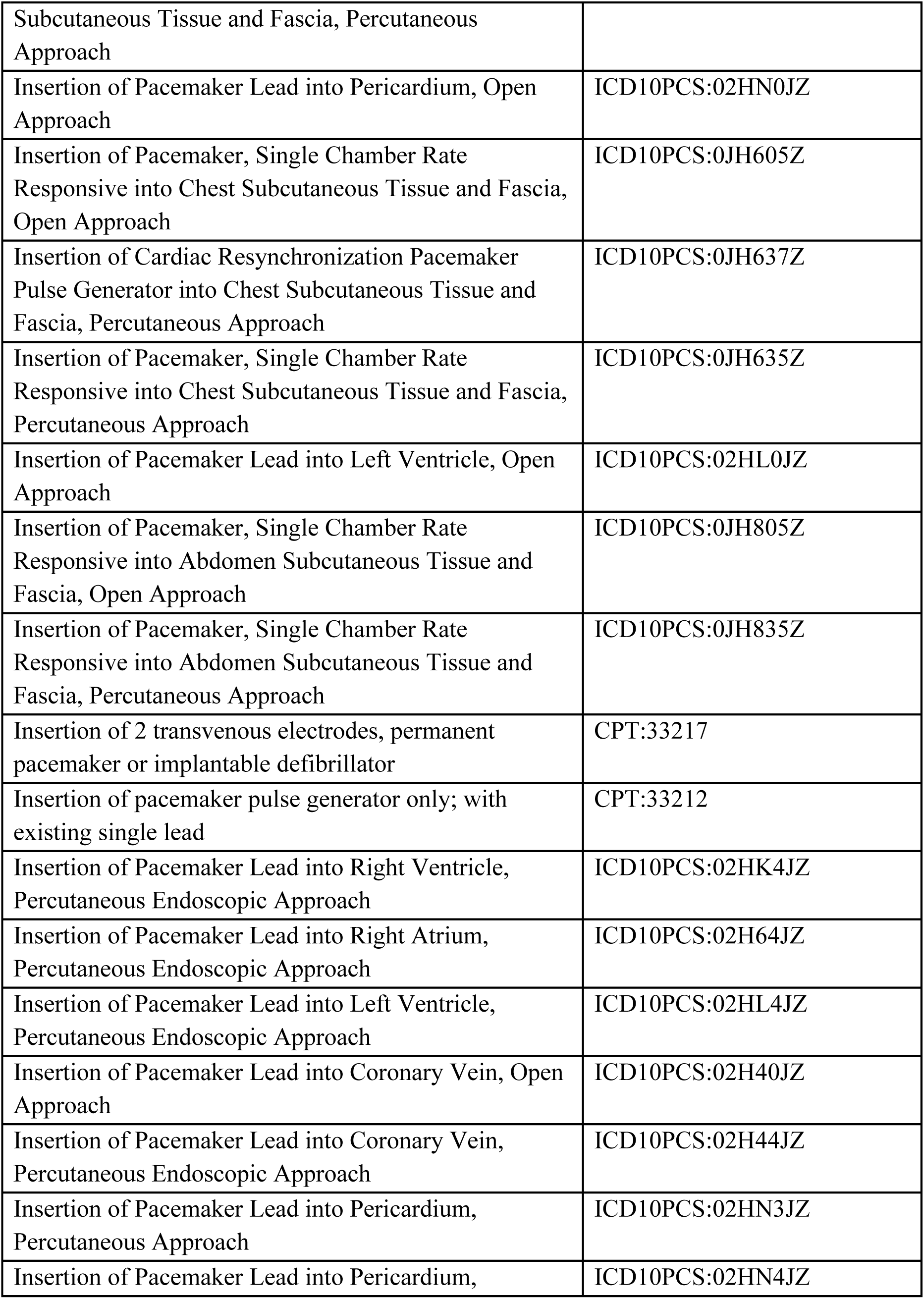
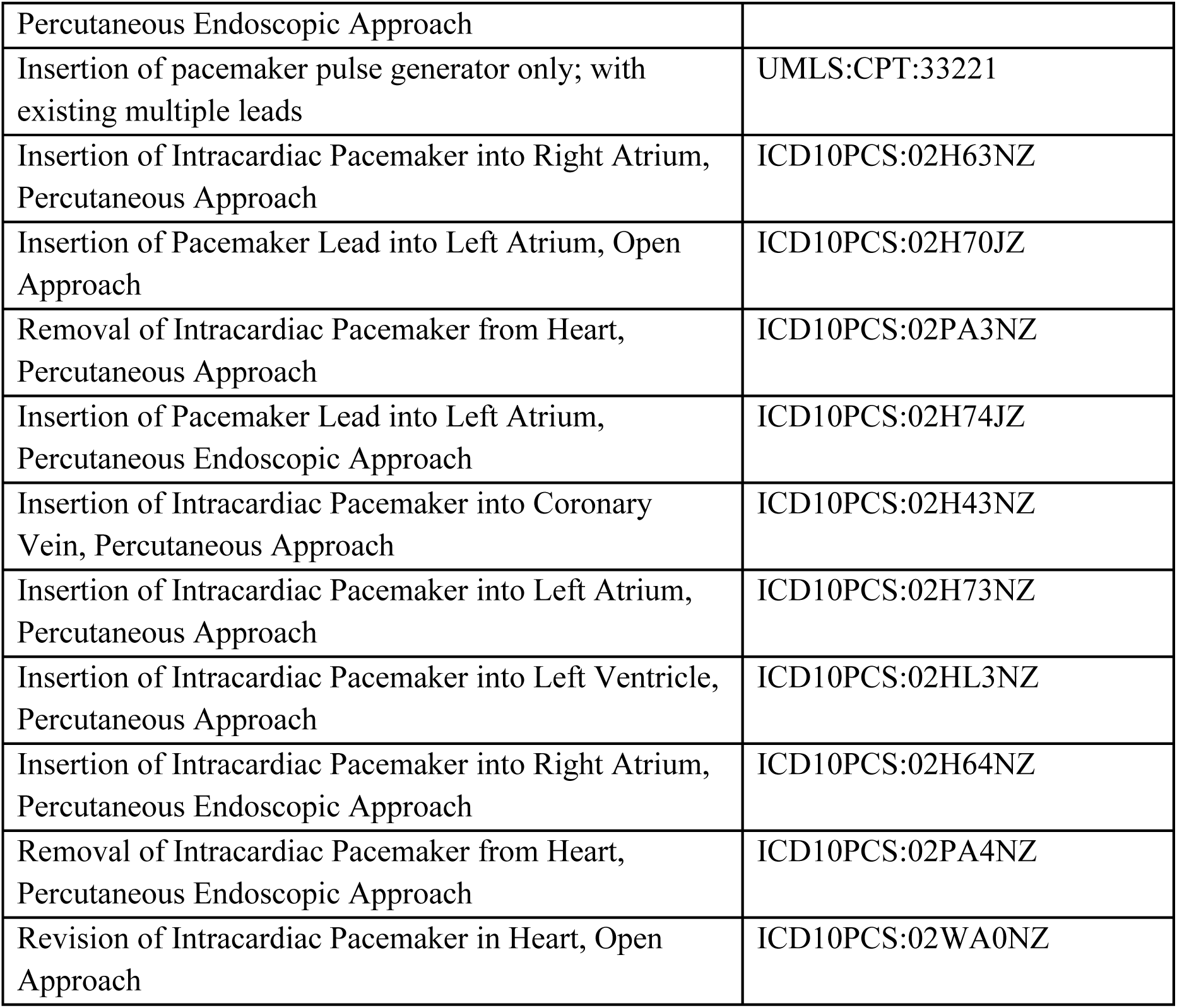
The terms in this group occurred between Jan 1, 2013, and Dec 31, 2023. Patients must have any of the following:

